# Bedside execution, not schedule mismatch: characterizing inpatient carbidopa-levodopa administration timing in Parkinson disease

**DOI:** 10.64898/2026.08.16.26360535

**Authors:** Jake Plagenz, Amy Lin, Tanya Harlow

## Abstract

**Background:** Timely carbidopa-levodopa administration is a recognized inpatient safety priority in Parkinson disease, and mistiming is common, but where in the medication-use process it arises is uncharacterized.

**Objectives:** To localize where inpatient mistiming arises and where to target intervention.

**Methods:** In a single-center retrospective analysis of hospitalized adults with Parkinson disease on home carbidopa-levodopa, each dose’s administration time was compared with the individualized home schedule. Mistiming was defined a priori as more than 15 minutes from the home time (Parkinson’s Foundation Hospital Care Standard 2). We characterized the deviation distribution, tested whether administrations tracked the schedule or the standard grid, and examined length-of-stay and readmission.

**Results:** Across 947 doses in 101 patients, ordering was accurate, yet 62.9% (596 of 947) missed the home time by more than 15 minutes and 99% of patients had at least one mistimed dose. Administrations tracked the individualized schedule almost exactly (Pearson r 0.98), not the standard grid: only 10% fell within 15 minutes of the default times, and the median dose sat 24 minutes from its home time but 76 from the nearest default. Deviation was symmetric drift (median absolute deviation 24 minutes; 16.5% beyond 60 minutes) and was not associated with length of stay (Spearman -0.11).

**Conclusions:** Mistiming reflected imprecise bedside execution, not ordering or a mismatch between fixed rounds and individualized regimens. These findings may point medication-safety efforts toward protecting bedside administration as complementary to redesigning orders.

## Introduction

Carbidopa-levodopa remains the foundation of symptomatic Parkinson disease treatment, and its short plasma half-life means that its motor benefit is tightly coupled to the timing of each dose. Patients with more advanced disease and motor fluctuations live within narrow windows between the medicated “on” state and the unmedicated “off” state, and many manage this at home with individualized schedules specified to the minute. Even modest deviations from that schedule can precipitate off periods, with loss of mobility, impaired swallowing and speech, falls, and distress.^1,2^

Hospitalization disrupts this finely tuned routine. Inpatient medication errors involving antiparkinsonian drugs are common and clinically consequential, and timely administration has been formalized as an explicit standard of inpatient care, the Parkinson’s Foundation Hospital Care Standard 2, which sets a 15-minute window around the patient’s home schedule.^3^ Prior work has documented high rates of mistimed and missed doses across inpatient and emergency settings and has associated deviations with prolonged length of stay and other adverse outcomes. ^1,4,5,6^ A parallel literature shows that targeted interventions can improve timing, including custom order entry that captures patient-specific times, multidisciplinary quality-improvement bundles, and pharmacy-led measures.^7,8,9^

What has not yet been definitively localized is where in the medication pathway the failure occurs at a given institution. The prevalence of mistiming and its association with worse outcomes are well documented, and interventions such as custom order entry can improve timing, but which point in the pathway fails, and therefore which lever applies, has rarely been examined directly. A common and intuitive assumption, reflected in much of the literature and in advocacy, is that fixed hospital medication rounds fail to accommodate the individualized, frequent schedules that Parkinson disease requires. If true, the corrective lever is order redesign, so that administrations follow patient-specific times rather than default rounds. If instead ordering is accurate and administrations already follow the individualized schedule but drifts at the bedside, the lever is execution rather than order design. These are distinct problems that require looking at where administrations actually fall relative to both the home schedule and the hospital’s standard passes.

We therefore conducted a single-center retrospective analysis of inpatient carbidopa-levodopa administration in patients with Parkinson disease. Our aims were to quantify mistiming against the 15-minute standard, to determine whether mistiming reflected incorrect ordering, clustering at fixed hospital passes, or drifts around the correct individualized time, and to characterize the severity distribution and explore outcome associations, in order to identify where intervention should be prioritized. We framed the study as a characterization and targeting analysis rather than an intervention trial.

## Methods

### Context and design

This was a single-center retrospective observational analysis conducted within the Sanford Health system. On September 16, 2025, the Sanford Health Institutional Review Board determined that the project did not constitute human subjects research and that IRB review and approval were not required. All patient data were de-identified for analysis.

### Cohort

We included hospitalized adults with a diagnosis of Parkinson disease who were taking a home carbidopa-levodopa regimen. Patients with a diagnosis of Parkinson disease were identified using SlicerDicer (Epic), and admissions from January 2021 through August 2025 were requested. Admissions during which no inpatient carbidopa-levodopa dose was administered were excluded.

### Data sources and reconciliation

The home regimen, including drug, dose, formulation, and the specific scheduled times, was obtained from the admission medication reconciliation and the documented home medication list. Inpatient administrations were obtained from the electronic medication administration record. Each administered inpatient dose was matched to its intended home dose and scheduled time.

### Ordering accuracy

Reconciliation accuracy at admission was assessed as agreement between the ordered inpatient regimen and the home regimen on dose, formulation, and frequency. This measure was used to localize the timing problem relative to prescribing and is reported as a single descriptor rather than a primary outcome. It does not capture whether contraindicated co-medications were ordered, a limitation addressed below.

### Timing measure and benchmark

For each dose, the signed deviation was calculated as the actual administration time minus the scheduled home time, in minutes, on a 24-hour circular scale so that times near midnight were handled correctly. Mistiming was defined as an absolute deviation greater than 15 minutes, consistent with Parkinson’s Foundation Hospital Care Standard 2. Magnitude was binned as 15 minutes or less, 16 to 30, 31 to 60, and more than 60 minutes, and direction was classified as early or late.

Order scheduling and administration-time clustering. At this institution, inpatient carbidopa-levodopa orders default to standard, frequency-specific administration times (two times daily at 09:00 and 21:00; three times at 09:00, 15:00, and 21:00; four times at 09:00, 13:00, 17:00, and 21:00; five times at 08:00, 11:00, 14:00, 18:00, and 21:00), and a pharmacist can override these to the patient’s home times using the schedule-adjustment tool or administration comments that propagate to the medication administration record. To distinguish drift around the individualized schedule from a default to these standard times, we (1) correlated actual with scheduled administration times across all doses, (2) computed, for each dose, the distance from the actual time to its own scheduled home time and to the nearest standard default time for that patient’s dosing frequency, and (3) compared the proportion of actual and of scheduled times falling within 15 minutes of the default grid. Patients dosing six or more times daily, for whom no standard grid is defined, were retained in the deviation analyses but excluded from the frequency-default comparison. We additionally summarized deviation by scheduled time of day to identify whether mistiming concentrated at particular administration windows.

### Outcomes

Length of stay (admission to discharge, in days) and 30-day readmission were extracted from the record and treated as exploratory, hypothesis-generating outcomes. To avoid conflating exposure with opportunity, the per-patient exposure was the mistiming rate (the proportion of that patient’s doses that were mistimed) rather than a raw count of mistimed doses, because longer stays mechanically accrue more doses and more opportunities for mistiming.

### Statistical analysis

Because doses are nested within patients and are not independent, we addressed clustering by reporting both dose-level and patient-level summaries and by conducting all outcome analyses at the patient level, using the per-patient mistiming rate rather than individual doses as the unit of analysis; we did not compute dose-level significance tests that would treat the 947 doses as independent observations. Continuous variables are summarized as median and interquartile range. Associations used Spearman correlation, and 30-day readmission was compared across mistiming-rate tertiles using Fisher’s exact test. Analyses were performed in Python 3.12.3 (pandas 3.0.2, NumPy 2.4.4, and SciPy 1.17.1), and figures were produced with Matplotlib 3.10.8. Two-sided p values below 0.05 were considered significant. The study was powered to characterize the timing distribution and was not powered to detect outcome differences; outcome analyses are accordingly interpreted as exploratory.

## Results

### Cohort

The cohort comprised 101 patients contributing 947 inpatient carbidopa-levodopa doses. The median number of doses per patient was 10 (IQR 5 to 12, range 1 to 24). The median length of stay was 3 days (IQR 1 to 5, range 0 to 16), and 21 of 101 patients (20.8 percent) were readmitted within 30 days. Of 947 doses, 931 had both an actual and a scheduled time available and entered the timing analysis; 16 carried an actual time without a recorded scheduled comparator and 2 were empty rows, and these were excluded from deviation and clustering analyses. Recorded formulations were predominantly immediate-release, with smaller numbers of extended- and controlled-release preparations (Table 1).

**Table 1.**
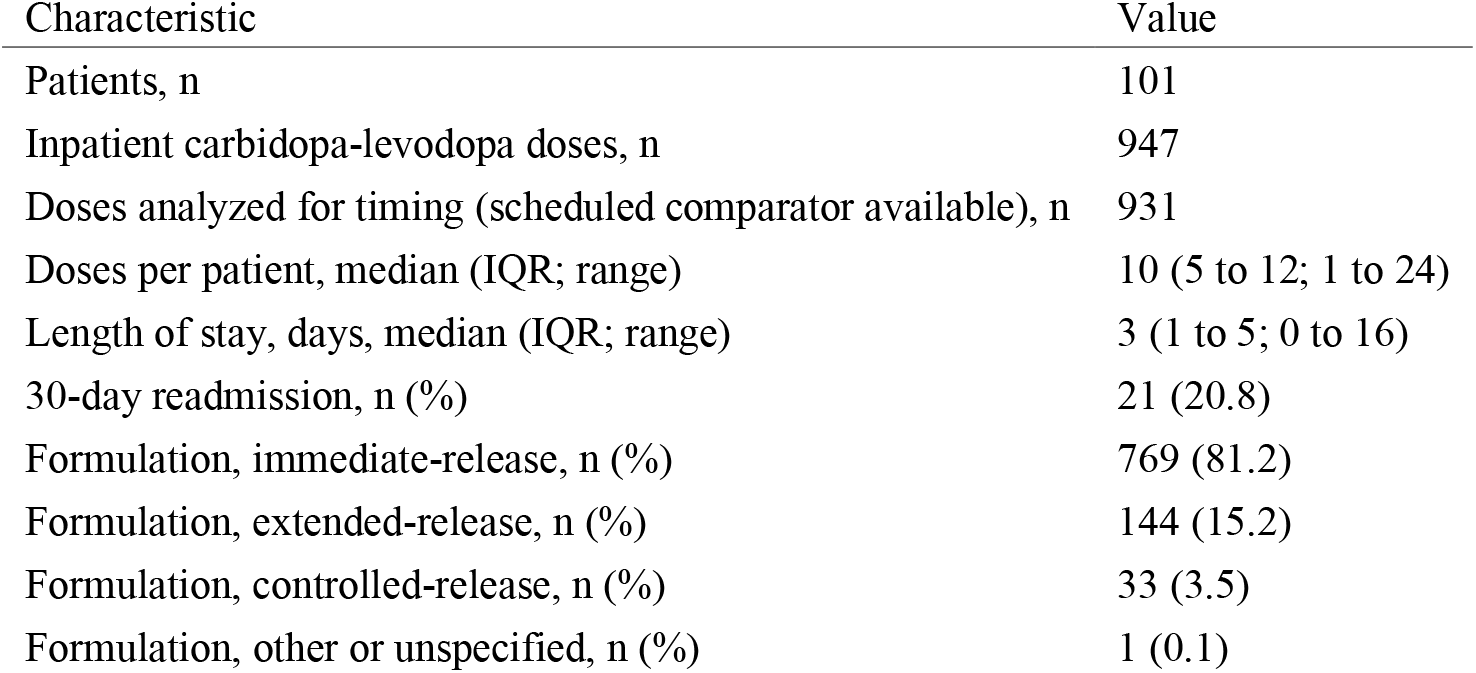
Cohort and dose characteristics.

| Characteristic | Value |
| --- | --- |
| Patients, n | 101 |
| Inpatient carbidopa-levodopa doses, n | 947 |
| Doses analyzed for timing (scheduled comparator available), n | 931 |
| Doses per patient, median (IQR; range) | 10 (5 to 12; 1 to 24) |
| Length of stay, days, median (IQR; range) | 3 (1 to 5; 0 to 16) |
| 30-day readmission, n (%) | 21 (20.8) |
| Formulation, immediate-release, n (%) | 769 (81.2) |
| Formulation, extended-release, n (%) | 144 (15.2) |
| Formulation, controlled-release, n (%) | 33 (3.5) |
| Formulation, other or unspecified, n (%) | 1 (0.1) |

### Ordering was accurate; timing was not

Ordering and reconciliation at admission were accurate for dose, formulation, and frequency in all 101 patients, which localizes the problem downstream of prescribing. Nonetheless, 596 of 947 doses (62.9 percent) were administered more than 15 minutes from the patient’s home schedule, and 100 of 101 patients (99 percent) experienced at least one mistimed dose.

### Administrations tracked the individualized schedule, not the standard grid

Actual administration times tracked the scheduled home times almost exactly (Pearson r 0.98; Spearman 0.98) (Figure 1A). The home schedules were themselves highly individualized: across patients dosing two to five times daily, home times sat a median of 90 minutes from the standard default grid for their frequency, and 98 percent of these patients had a home schedule more than 15 minutes from the default grid. Administrations did not concentrate at the standard default times: among the 871 doses in patients with a defined default grid, the median dose was administered 24 minutes from its own scheduled home time but 76 minutes from the nearest standard default time, only 10 percent fell within 15 minutes of the default grid, and 71 percent were closer to their own home time than to any default time (Figure 1B). Because administrations sat far from the default grid rather than near it, the individualized times were being captured on the medication administration record rather than left at the defaults. Mistiming in this cohort therefore did not reflect a substitution of the standard grid for the individualized regimen.

**Figure 1.**
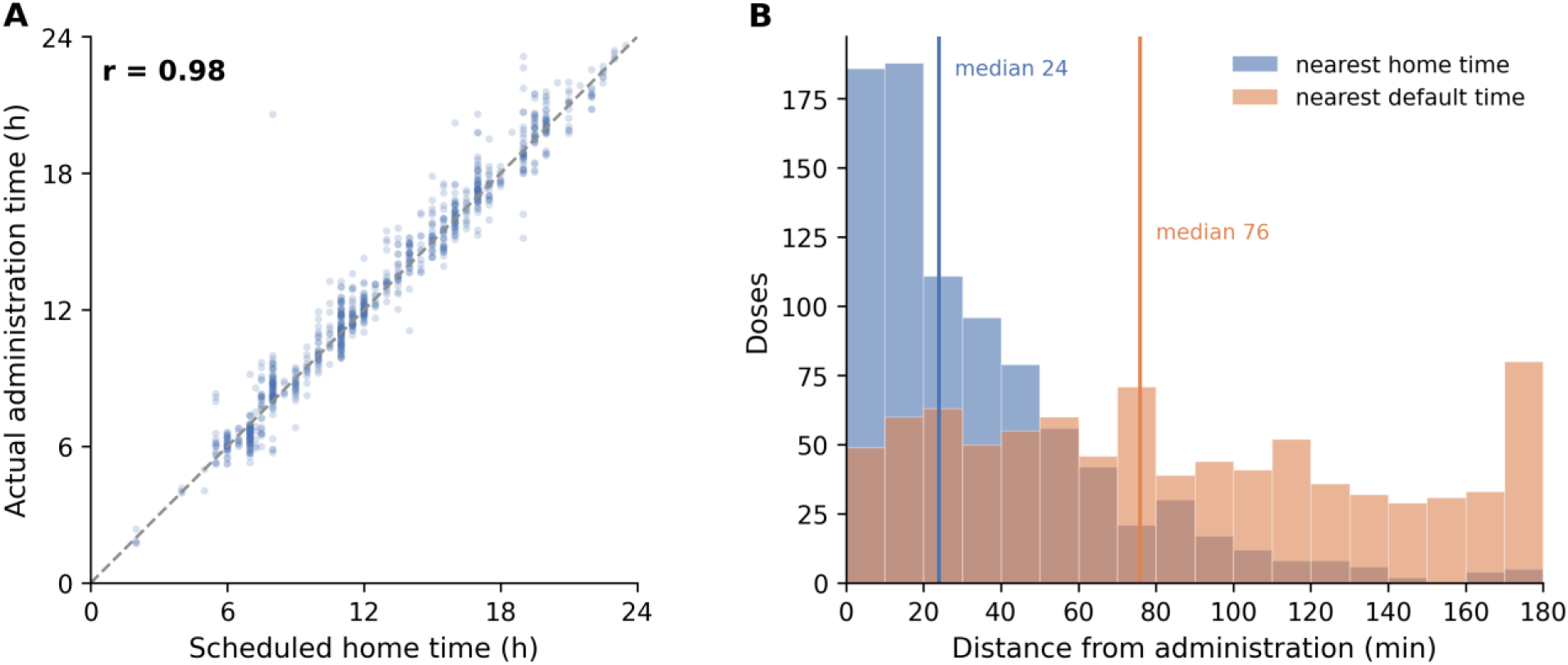
Administrations track the individualized schedule, not the standard default grid. (A) Scheduled home time versus actual administration time for the 931 analyzable doses, with the line of identity; Pearson r 0.98. (B) For the 871 doses in patients dosing two to five times daily, the distance from each administration to its nearest scheduled home time (median 24 minutes) and to the nearest standard default time for that dosing frequency (median 76 minutes); administrations cluster near the home time and sit far from the default grid.

### The mistiming drifts around the correct target

The signed deviation distribution was centered near zero (median +3 minutes) with substantial spread (Figure 2). The median absolute deviation was 24 minutes (IQR 11 to 48). By magnitude, 36.7 percent of doses were within 15 minutes, 20.5 percent were 16 to 30 minutes off, 26.2 percent were 31 to 60 minutes off, and 16.5 percent were more than 60 minutes off. Among mistimed doses, 58 percent were late and 42 percent were early, a modest late predominance.

**Figure 2.**
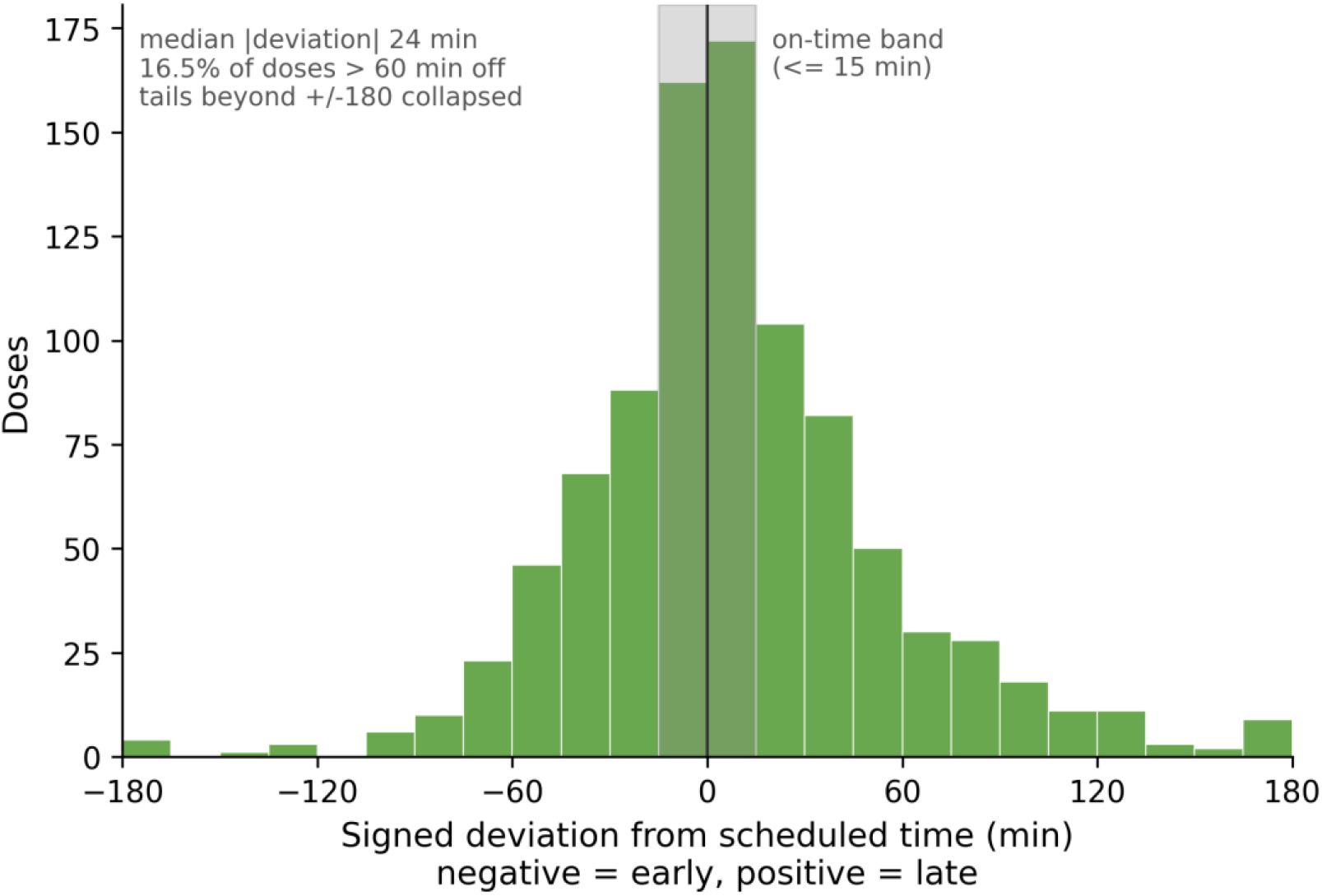
The mistiming drifts around the correct target. Signed deviation from the scheduled home time (negative early, positive late) for the 931 analyzable doses; the on-time band (15 minutes or less) is shaded. Median absolute deviation 24 minutes; 16.5 percent of doses more than 60 minutes off.

### The drift was pervasive across administration times

Median absolute deviation ranged narrowly from 20 to 26 minutes across time-of-day windows, and more than half of doses were mistimed in every window (Table 2). The evening window (1700 to 2059) showed the highest proportion of doses more than 60 minutes off (23 percent) and the highest proportion administered late (42 percent), while the midday window (0900 to 1259) was least affected. The earliest scheduled dose of the day was mistimed slightly more often than later doses (68 versus 61 percent). No window was spared, consistent with a systemic rather than a localized problem.

**Table 2.** Timing results and exploratory outcome associations.

| Measure | Value |
| --- | --- |
| **Overall timing** |  |
| Mistimed (more than 15 min from home schedule), n (%) of 947 | 596 (62.9) |
| Patients with at least one mistimed dose, n (%) | 100 (99.0) |
| Median signed deviation, min | +3 |
| Median absolute deviation, min (IQR) | 24 (11 to 48) |
| **Deviation magnitude (n = 931)** |  |
| 15 min or less (on time), n (%) | 342 (36.7) |
| 16 to 30 min, n (%) | 191 (20.5) |
| 31 to 60 min, n (%) | 244 (26.2) |
| More than 60 min, n (%) | 154 (16.5) |
| Late vs early among mistimed doses, % | 58 vs 42 |
| **Exploratory outcomes (patient level, n = 101)** |  |
| Per-patient mistiming rate vs length of stay | Spearman -0.11 (p 0.26) |
| Dose count vs length of stay | Spearman 0.80 (p < 0.001) |
| Readmission by mistiming-rate tertile (low, mid, high) | 26%, 12%, 25% |

*Deviation by scheduled time of day (n = 931)*
| Window | Doses, n | Median abs. deviation, min | More than 15 min, % | More than 60 min, % | Late, % |
| --- | --- | --- | --- | --- | --- |
| Before 0900 | 243 | 25 | 67.5 | 16.9 | 35.8 |
| 0900 to 1259 | 265 | 20 | 57.0 | 12.8 | 33.6 |
| 1300 to 1659 | 198 | 26 | 64.6 | 15.2 | 39.4 |
| 1700 to 2059 | 189 | 26 | 65.1 | 23.3 | 41.8 |
| 2100 or later | 36 | 22 | 63.9 | 13.9 | 19.4 |

Timing analyses use the 931 doses with both a recorded administration time and a scheduled home comparator; 16 doses lacked a comparator and 2 empty rows were removed. Outcome analyses are exploratory and underpowered.

### Mistiming was not associated with outcomes

The per-patient mistiming rate was not associated with length of stay (Spearman -0.11, p 0.26), and 30-day readmission did not differ across mistiming-rate tertiles (26 percent, 12 percent, and 25 percent from lowest to highest tertile). Length of stay was instead strongly associated with dose count (Spearman 0.80, p < 0.001), confirming that a raw count of mistimed doses would have produced a spurious association driven by length of stay rather than by mistiming itself (Table 2). These outcome analyses were underpowered and are reported as exploratory.

## Discussion

In this single-center cohort, inpatient carbidopa-levodopa mistiming was common, affecting nearly two-thirds of doses and almost every patient, yet its source was not where it is often assumed to be. Ordering was accurate, and administrations tracked the individualized home schedule almost exactly rather than defaulting to fixed hospital passes. Therefore, an area of improvement is one of bedside execution precision: even with the correct drug scheduled at the correct individualized time, the administered dose drifted a median of 24 minutes, and roughly one in six doses landed more than an hour off. The principal finding is best summarized as a three-part elimination. The gap was not a knowledge or prescribing gap, because ordering was accurate. It was not a schedule-mismatch gap, because administrations followed the individualized schedule. It was a gap in the reliable execution of an already-correct plan at the bedside.

This reframing matters because it differs from frequently cited interventions. Custom order entry, which aligns orders with patient-specific times, addresses schedule mismatch and is an important tool at institutions where that mismatch exists; in the largest such study it meaningfully improved on-time administration.^8^ At our institution, orders default to standard frequency-based times, but pharmacists individualize them to the home schedule through the schedule-adjustment tool or administration comments; the finding that administrations sat a median of 76 minutes from the default grid, and only 10 percent within 15 minutes of it, indicates that this individualization was in fact reaching the medication administration record for most patients. This is consistent with an external inpatient program in which pharmacist-led medication histories raised the proportion of orders matching the home regimen from 40 to 89 percent.^10^ In other words, the schedule-mismatch problem that custom order entry solves was already being solved upstream here, yet nearly two-thirds of doses still missed the window. Order-level scheduling remains necessary but not sufficient, and where the individualized times are already captured the residual drift localizes downstream, to execution at the bedside, leaving order redesign little additional room to help.

The lever here is protecting execution at the bedside: designating carbidopa-levodopa as time-critical so that it is treated with the urgency afforded to insulin and antibiotics, prioritizing these administrations within nursing workflow, and enabling bedside availability where safe. ^9,11^ The presence of drift across every administration window, rather than confined to a single understaffed shift, argues for a systemic time-critical approach in addition to a narrow staffing fix, although the modestly worse evening performance is a reasonable place to concentrate first.

Our outcome analysis was null, and the reason is instructive. Prior work has associated timing deviations with longer length of stay.^1^ We found no association between the per-patient mistiming rate and length of stay, while dose count was strongly associated with length of stay. Because sicker patients with longer admissions receive more doses and thereby accrue more opportunities for mistiming, an analysis that uses a raw count of mistimed doses as the exposure will tend to manufacture an association that reflects length of stay rather than timing. Using a per-patient rate removes that artifact, and the association disappears in our data. We interpret this cautiously given the modest sample and single-center design, but it is a concrete methodological caution for future outcome analyses and argues that they should model exposure as a rate and account for the competing relationship between dose count and length of stay.

### Limitations

This was a single-center, retrospective analysis, and the finding that administrations closely tracked home schedules may not generalize: an institution with this degree of schedule fidelity may already have effective order capture, while a site where orders default to fixed times may face genuine schedule mismatch, for which order redesign rather than bedside measures would be the lever. The transferable point is therefore not that execution is the dominant failure everywhere, but that the locus of failure should be diagnosed rather than assumed, and the simple comparison used here, of administered times against both the individualized schedule and the fixed passes, offers a low-cost way to make that diagnosis before an intervention is chosen. We did not extract the exact scheduled times encoded in each inpatient order, so we infer rather than directly confirm that the individualized times were captured at the order level; that administrations sat far from the standard default grid, together with the local pharmacist workflow of overriding defaults to the home schedule, makes order-level capture the most supported explanation, though a residual contribution of bedside correction cannot be excluded. This individualization is pharmacist-dependent and not enforced, so schedule fidelity may vary by pharmacist and by site, which bears on generalizability. The outcome analyses were underpowered, and the null associations should not be read as evidence of no effect.

Reconciliation accuracy was assessed on dose, formulation, and frequency and did not capture whether contraindicated antidopaminergic medications were co-ordered, so the ordering-accuracy statement is narrower than a complete prescribing-safety assessment. Formulation was captured at the level of release type (immediate-, extended-, or controlled-release) rather than specific product, limiting product-level inference. Retrospective records do not capture legitimate reasons for delay such as nil-per-os status, procedures, patient absence from the unit, or refusal, which a prospective design would document. Finally, deviation was measured against the home schedule as recorded, and errors in that reference would propagate to the deviation estimate.

### Future directions

These findings motivate prospective, multi-site evaluation to test whether the execution-precision pattern holds across institutions, and targeted implementation studies of bedside time-critical measures with adequately powered outcome assessment using competing-risk methods for length of stay.

## Conclusion

Inpatient carbidopa-levodopa mistiming in this cohort was a problem of bedside execution, not of ordering or of a mismatch between fixed hospital rounds and individualized regimens. Efforts to improve inpatient Parkinson disease care should prioritize protecting timely administration at the bedside and should be directed by where the drift is most severe.

## Acknowledgments

The authors thank Elizabeth Gau, PharmD, BCCCP, for clarifying the inpatient carbidopa-levodopa order-entry and scheduling workflow, and Alexis Meyer, for acting as project manager. During the preparation of this work, the authors used Claude (Anthropic) for copy-editing and formatting of manuscript materials; the tool was not used for study conception, data analysis, or authorship, and the authors reviewed and edited all content and take full responsibility for the article.

## Author Contributions

Jake Plagenz: conceptualization, methodology, software, formal analysis, investigation, data curation, visualization, writing (original draft), and writing (review and editing). Amy Lin: investigation, data curation, writing (original draft), and writing (review and editing). Tanya Harlow: supervision, writing (original draft), and writing (review and editing). All authors reviewed and approved the final manuscript.

## Ethics Statement

On September 16, 2025, the Sanford Health Institutional Review Board determined that this project did not constitute human subjects research (STUDY00004067) and did not require review or approval. Informed patient consent was not required because the analysis used only de-identified retrospective data.

## Data Availability Statement

The de-identified data supporting the findings of this study are available from the corresponding author upon reasonable request. The data are not publicly available because of institutional privacy and data-governance restrictions.

